# Clinical Implementation of an AI Algorithm for Substance Misuse Screening in Hospitalized Adults

**DOI:** 10.1101/2025.11.17.25340323

**Authors:** Juan C. Rojas, Cara Joyce, Talar W. Markossian, Vaishvik Chaudhari, Mia R. McClintic, Fatima Castro, AJ Fairgrieve, Dmitriy Dligach, Madeline K. Oguss, Matthew M. Churpek, Jenna Nikolaides, Majid Afshar

## Abstract

**Importance:** Manual inpatient screening for substance misuse is labor-intensive and inconsistently applied. Evaluation of artificial intelligence (AI)–assisted screening during clinical implementation is needed to determine clinical and economic performance.

**Objective:** To assess whether an AI-based screening program with the Substance Misuse Algorithm for Referral to Treatment Using Artificial Intelligence (SMART-AI) maintained delivery of addiction-related services compared with manual screening and to evaluate readmissions and costs.

**Design, Setting, and Participants:** Prospective, quasi-experimental pre–post study at a large academic medical center in Chicago, Illinois between 2022 and 2025. The pre-implementation period (manual screening) included 31,432 hospitalizations, and the post-implementation period with AI augmentation (SMART-AI) included 33,564.

**Interventions/Exposures:** During the post-implementation period, SMART-AI screened clinical documentation within 24 hours of admission to identify patients at-risk for a substance use disorder and notified the Substance Use Intervention Team. In the pre-implementation period, screening relied on manual processes, with nurses and social workers screening with standardized questionnaires.

**Main Outcomes and Measures:** The primary outcome was receipt of ≥1 addiction-related service (initiation or adjustment of medication for alcohol or opioid use disorder; brief intervention/motivational interviewing; naloxone dispensing; or a completed addiction medicine consultation). The prespecified noninferiority margin was −0.5 percentage points (1-sided α = 0.025). Secondary outcomes included 6-month readmission, discharge against medical advice, and program costs.

**Results:** Addiction-related services were received in 1,189 of 31,432 hospitalizations (3.8%) during manual screening and 1,144 of 33,564 (3.4%) during SMART-AI (difference, −0.4 percentage points; 95% CI, −0.7 to −0.1; P = 0.20). The lower limit of the confidence interval was below the noninferiority margin, so noninferiority was not achieved. Six-month readmissions across all hospitalizations occurred in 9,586 patients (30.5%) in the manual period and 10,244 patients (30.5%) in the SMART-AI period (P = 0.95), and discharge against medical advice did not differ (1.3% v. 1.1%). Among patients who received a SUIT intervention (n = 2,296), 6-month readmission occurred in 41.3% (485/1,175) during usual care versus 37.0% (415/1,121) during SMART-AI (odds ratio: 0.86, 95% CI: 0.73-1.03, p=0.10). Program costs over a 1-year period were $6,166.71 lower after SMART-AI automation.

**Conclusions and Relevance:** AI-assisted screening did not meet the prespecified non-inferiority criterion for maintaining service delivery, but it was associated with maintaining secondary outcomes among patients screened for substance use disorder with lower program costs. Findings support the feasibility and potential value of automated screening at scale.

Trial RegistrationClinicalTrials.gov Identifier NCT03833804

## INTRODUCTION

Unhealthy use of alcohol, opioids, and other illicit or non-prescribed drugs now accounts for more hospital encounters in the United States than heart disease or respiratory failure, with drug overdose deaths reaching a record high of 93,000 in 2020^1,2^. Among hospitalized adults, the prevalence of substance misuse is estimated at 15%–25%, yet fewer than half of the cases are identified through routine screening, and less than one-quarter of those receive evidence-based treatment prior to discharge^3,4^. These care gaps directly lead to preventable morbidity, mortality, and rehospitalization.

Screening, Brief Intervention, and Referral to Treatment (SBIRT) programs remain the recommended standard of care, but their reliance on nurses and social workers to administer tools such as the Tobacco, Alcohol, Prescription medication, and other Substance use (TAPS) imposes a human labor-intensive, non-reimbursable workflow^5–7^. These programs are inconsistently implemented, vulnerable to staffing shortages, and were further strained during the COVID-19 pandemic^8^. Even in health systems that have invested in SBIRT infrastructure, the operational burden has limited scalability and sustainability, driving interest in automated solutions that can identify at-risk patients using electronic health record (EHR) data while preserving finite clinical resources^9,10^.

Recent advances in artificial intelligence (AI) have enabled transforming free-text clinical notes into structured inputs for AI classifiers. The Substance Misuse Algorithm for Referral to Treatment Using Artificial Intelligence (SMART-AI) was developed on a hospitalized cohort of adults with a 3.5% prevalence of substance misuse^11,12^. This multilabel deep learning model detects alcohol, opioid, and non-opioid misuse at hospital admission from EHR narrative documentation. In validation, SMART-AI showed strong performance across substance categories and maintained equitable predictions across racial groups^11,13^. By enabling early and unbiased detection of substance use disorders, the model provides a scalable, low-friction approach to increasing access to evidence-based care in hospitals. A prior implementation study demonstrated that AI-driven screening for opioid use disorder was associated with a reduction in 30-day readmissions compared with manual screening^14^.

However, studies evaluating real-world implementation of AI-based screening for substance use disorders beyond opioid use disorder—such as alcohol use disorder—remain absent from the current literature. Evaluating comprehensive models capable of detecting multiple use disorders, as well as polysubstance use, is essential for determining the feasibility and scalability of AI-assisted tools to enhance addiction medicine care.

In this study, we evaluated the real-world impact of SMART-AI by replacing manual SBIRT with: (1) nightly batch processing of admission notes, (2) automated classification of alcohol, opioid, and non-opioid misuse, and (3) secure chat notifications to clinicians on screen positives from the AI algorithm^11^. Our approach implemented SMART-AI through EHR-embedded Secure Chat, delivering results to the Substance Use Intervention Team (SUIT) via human messaging for multiple substance use types, allowing for a more comprehensive screening tool that can also capture polysubstance use^15^. This unobtrusive workflow acts as a clinical “nudge,” linking AI recommendations with bedside decisions. Over a 12-month period, we assessed changes in delivery of a composite intervention (where the patient received at least one of the following: medication-assisted treatment, brief intervention/motivational interviewing, naloxone dispensing, or a completed addiction medicine consult) and conducted a parallel cost-effectiveness analysis.

## METHODS

We conducted a prospective, quasi-experimental pre-post study at Rush University Medical Center (RUMC) in Chicago to evaluate an AI-assisted Natural Language Processing (NLP) screening program for unhealthy substance use among hospitalized adults. The 24-month study included a pre-intervention period of manual screening, spanning 12 months from September 19, 2022, to September 19, 2023. This was followed by a 12-month post-intervention period (during which the automated screening tool was deployed) from 20 September 2023 to 19 September 2024, and a 6-month follow-up period ending on 24 March 2025 for evaluation of secondary outcomes. It was approved by the Rush Institutional Review Board (ORA# 22040503) and registered at ClinicalTrials.gov (NCT03833804). Eligible participants included all adult inpatients over 18 years old admitted to medical and surgical units during the study period. The full clinical trial protocol was previously published^16^.

### Interventions

#### Pre-intervention Period: Manual Screening

During the manual screening phase, all patients underwent routine screening in accordance with the hospital’s SBIRT protocol^9^. Nurses administered a two-question universal screening tool to assess recent alcohol and drug use. Patients who screened positive were referred to floor social workers, who conducted full screenings using the Alcohol Use Disorders Identification Test (AUDIT) and/or Drug Abuse Screening Tool (DAST)^17,18^. Based on assessment scores and discussions with the patient, social workers could prompt a consultation with the hospital’s addiction medicine consult service (SUIT). Alternatively, primary teams were able to consult the SUIT directly, at which time, AUDITs and DASTs were performed by the SUIT themselves. SUIT, comprising providers from addiction medicine, psychiatry, social work, and pharmacy, offers interventions such as motivational interviewing, medication-assisted treatment (MAT), naloxone distribution, and outpatient addiction treatment referrals^19^.

#### Post-intervention Period: AI-Assisted Screening

During the automated screening phase, manual nurse and social work screenings were replaced by an AI-based tool called SMART-AI^11^. It was a validated convolutional neural network model trained to identify unhealthy substance use based on unstructured clinical notes recorded within the first 24 hours of hospitalization. The fully trained model, methods, and prior work are publicly available at https://git.doit.wisc.edu/smph-public/dom/uw-icu-data-science-lab-public/smart-ai^20^. Screen-positive patients were flagged in a daily email report sent to the study team at 8:00 AM. The SUIT team reviewed charts for hospitalized patients and, when appropriate, notified the primary team via Epic Secure Chat to recommend a SUIT consult. Patients unsuitable for consultation (e.g., medical contraindications, existing psychiatry care, or false-positive screens) received no follow-up (**Figure 1**). The primary team then decided whether to place a SUIT consult order. This workflow enabled early intervention consistent with prior SBIRT protocols.

**Figure 1.**
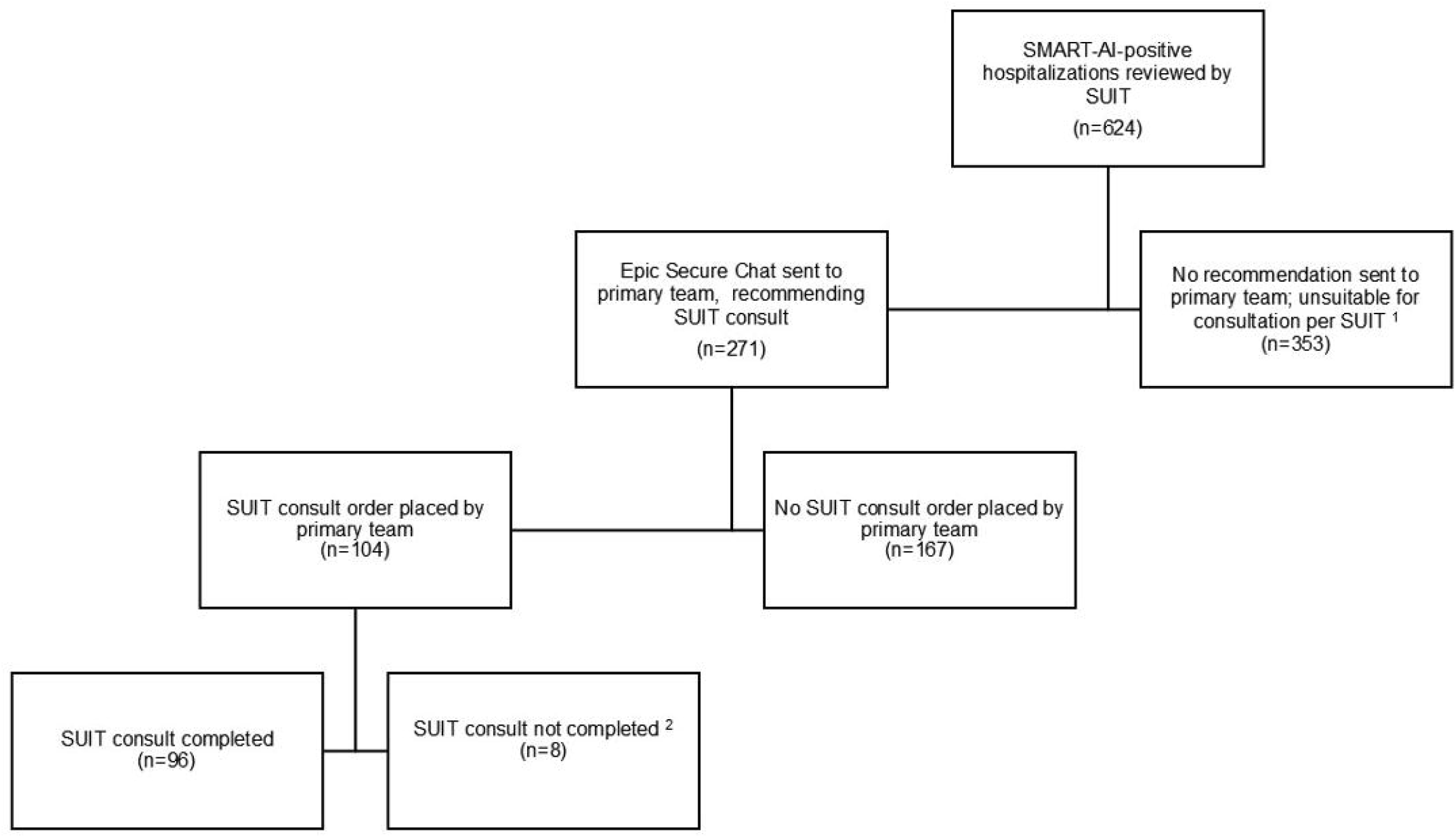
Flow Diagram of AI-Assisted Workflow During the SMART-AI Post-Implementation Period. Flow diagram of patient hospitalizations that screened positive by SMART-AI and were subsequently reviewed for real-time follow up during the post-implementation period. 1 SUIT did not prompt follow-up if, based on their chart review, consult would not be appropriate at that time. Reasons included deferral to psychiatry service (n=94), medical contraindications (n=38), deemed false positive (n=55), patient already declined (n=11), and other (n=155). 2 SUIT consult ordered but not completed due to various circumstances (e.g., patient left hospital before consult, consult deferred until after discharge).

#### Outcomes

The primary outcome was the proportion of hospitalizations in which at least one part of the composite intervention was delivered: (1) Medication-assisted treatment; (2) brief intervention/motivational interviewing; (3) naloxone dispensing; or (4) a completed addiction medicine consult. Secondary outcomes included all-cause hospital readmission within six months of discharge (overall and among those who received any component of the composite outcome), time from admission to SUIT consultation, and discharge against medical advice. We also examined each individual component of the composite intervention as an exploratory outcome.

#### Data Collection

Clinical data were collected monthly from the Rush University research data warehouse, linked to the EHR system (Epic Systems, Verona, WI). Documentation templates and flow sheets were updated before the study began to standardize recording screening methods, consultation reasons, intervention types, and patient outcomes (**eFigure1**). In addition to EHR data, clinical outcomes were tracked in real-time by monitoring each EHR chat nudge and its associated action, whether it prompted a consultation or not. For each nudge, the research team documented the clinical team’s response and, when relevant, the reason for declining a consultation. These data were systematically recorded using a REDCap (Research Electronic Data Capture) instrument^21^.

#### Statistical Analysis

We used descriptive statistics to summarize patient demographics and clinical characteristics by study period^22,23^. Based on 2020 data from RUMC, with an average of 2,400 hospitalizations and 94 SUIT consults per month (3.9% consult rate), we previously estimated that 60,000 total hospitalizations (30,000 per period) would provide 82% power at α = 0.025 (one-sided) to detect non-inferiority within a 0.5% margin between pre- and post-intervention periods for the primary outcome^16^. This margin, determined by addiction medicine and implementation experts, reflected a clinically acceptable tradeoff between impact and scalability rather than historical benchmarks. Multivariable logistic regression was used to compare the composite outcome between the pre- and post-intervention periods, with pre-specified covariate adjustment for age, sex, race, ethnicity, insurance, and hospital unit^16^. Secondary outcomes—six-month readmission and discharge against medical advice—were analyzed using logistic or Poisson regression with the same covariates^24^. Patients discharged to hospice care or who died in the hospital were excluded from readmission analyses. Analyses were conducted in R v4.5.0 (RStudio).

#### Cost-Effectiveness Analysis

We calculated the incremental cost-effectiveness ratio (ICER) as the cost per unit difference in effectiveness between automated and manual programs^25^. The analysis adopted a health system perspective over a 2-year horizon (12 months pre- and post-intervention). Program costs included fixed setup and variable monthly administrative costs, while hospitalization costs were derived from health system financial records and encompassed both direct and indirect billing costs. All costs were adjusted to 2024 U.S. dollars using the Centers for Medicare and Medicaid Services (CMS) Hospital Care Price Index^26,27^. Descriptive statistics summarized program, hospitalization, and readmission costs. Post-estimation models combined costs and effectiveness data to compute ICERs, with 95% confidence intervals estimated from 500 bootstrap samples. Sensitivity analyses varied program costs, outcomes, and sample definitions. Analyses were conducted using Stata v18 (College Station, TX).

## RESULTS

### Study Population

A total of 64,996 hospitalizations occurred during the 24-month study period, comprising 31,432 during the manual pre-period and 33,564 in the post-period with SMART-AI augmentation. Baseline characteristics were well balanced between groups (**Table 1**). The mean age was 56 years (SD 19) in both periods. Sex distribution was similar (58.7% vs 57.9% female), as were racial and ethnic compositions (Black 35.8% vs 35.1%, White 33.8% vs 33.4%, Hispanic 23.7% vs 24.3%). Inpatient admissions accounted for 77.1% of hospitalizations during usual care and 77.4% during the SMART-AI period; observation stays accounted for 22.9% and 22.6%, respectively. The emergency department was the primary admission source in both groups (48.8%). The median hospital stay was 3 days (IQR 2–6), and mean Elixhauser scores were similar (2.8 [SD 4.8] vs 2.9 [SD 4.9]). Medication exposures were comparable: opioids (36.0% vs 35.9%), naloxone (0.7% in both), and buprenorphine (1.5% vs 1.8%). Use of antidepressants, benzodiazepines, gabapentin, and antipsychotics was also similar. Most patients were discharged home (67.4% vs. 69.5%), and the proportion of patients leaving against medical advice was also similar (1.3% vs. 1.1%).

**Table 1.** Baseline patient characteristics.

| **Characteristic** | **Overall**<br>**(N = 64,996)** | **Usual Care**<br>**(N = 31,432)** | **SMART-AI**<br>**(N = 33,564)** |
| --- | --- | --- | --- |
| Age, years |  |  |  |
| Mean (standard deviation) | 56 (19) | 56 (19) | 56 (19) |
| Median (interquartile range) | 59 (40-71) | 58 (40-71) | 59 (40-71) |
| Sex, No. (%) |  |  |  |
| Female | 37,879 (58.3) | 18,458 (58.7) | 19,421 (57.9) |
| Male | 27,117 (41.7) | 12,974 (41.3) | 14,143 (42.1) |
| Race/ethnicity, No. (%) |  |  |  |
| White | 21,621 (33.6) | 10,524 (33.8) | 11,097 (33.4) |
| Black | 22,789 (35.4) | 11,139 (35.8) | 11,650 (35.1) |
| Hispanic | 15,424 (24.0) | 7,363 (23.7) | 8,061 (24.3) |
| Asian | 2,111 (3.3) | 1,047 (3.4) | 1,064 (3.2) |
| Other | 2,376 (3.7) | 1,058 (3.4) | 1,318 (4.0) |
| Unknown | 675 | 301 | 374 |
| Patient class, No. (%) |  |  |  |
| Inpatient | 50,223 (77.3) | 24,231 (77.1) | 25,992 (77.4) |
| Observation | 14,773 (22.7) | 7,201 (22.9) | 7,572 (22.6) |
| Admission type, No. (%) |  |  |  |
| Elective | 28,405 (43.7) | 13,753 (43.8) | 14,652 (43.7) |
| Emergency | 36,591 (56.3) | 17,679 (56.2) | 18,912 (56.3) |
| Admission origin, No. (%) |  |  |  |
| Direct admission | 8,927 (13.7) | 4,343 (13.8) | 4,584 (13.7) |
| Emergency department | 31,720 (48.8) | 15,330 (48.8) | 16,390 (48.8) |
| Operating room | 15,469 (23.8) | 7,557 (24.0) | 7,912 (23.6) |
| Transfer center | 8,875 (13.7) | 4,201 (13.4) | 4,674 (13.9) |
| Unknown | 5 | 1 | 4 |
| Weekend admission, No. (%) | 11,728 (18.0) | 5,702 (18.1) | 6,026 (18.0) |
| Inpatient medications, No. (%) |  |  |  |
| Opioids | 23,350 (35.9) | 11,308 (36.0) | 12,042 (35.9) |
| Gabapentin | 15,103 (23.2) | 7,352 (23.4) | 7,751 (23.1) |
| Antidepressants | 13,939 (21.4) | 6,621 (21.1) | 7,318 (21.8) |
| Antipsychotics | 5,707 (8.8) | 2,784 (8.9) | 2,923 (8.7) |
| Benzodiazepines | 4,774 (7.3) | 2,663 (8.5) | 2,111 (6.3) |
| Dexmedetomidine | 2,091 (3.2) | 1,024 (3.3) | 1,067 (3.2) |
| Propofol | 2,138 (3.3) | 1,024 (3.3) | 1,114 (3.3) |
| Clonidine | 1,472 (2.3) | 730 (2.3) | 742 (2.2) |
| Buprenorphine | 1,074 (1.7) | 481 (1.5) | 593 (1.8) |
| Ketamine | 630 (1.0) | 303 (1.0) | 327 (1.0) |
| Buspirone | 629 (1.0) | 278 (0.9) | 351 (1.0) |
| Naloxone | 430 (0.7) | 208 (0.7) | 222 (0.7) |
| Stimulants | 399 (0.6) | 185 (0.6) | 214 (0.6) |
| Barbiturates | 345 (0.5) | 188 (0.6) | 157 (0.5) |
| Naltrexone | 315 (0.5) | 179 (0.6) | 136 (0.4) |
| Acamprosate | 265 (0.4) | 153 (0.5) | 112 (0.3) |

| Characteristic | Overall<br>(N = 64,996) | Usual Care<br>(N = 31,432) | SMART-AI<br>(N = 33,564) |
| --- | --- | --- | --- |
| Elixhauser comorbidity score, mean (SD) | 2.9 (4.8) | 2.8 (4.8) | 2.9 (4.9) |
| Unknown | 1,656 | 764 | 892 |
| Length of stay, days |  |  |  |
| Mean (standard deviation) | 5.0 (6.5) | 5.1 (6.5) | 4.9 (6.4) |
| Median (interquartile range) | 3 (2-6) | 3 (2-6) | 3 (2-6) |
| Insurance type, No. (%) |  |  |  |
| Medicare | 27,475 (44.4) | 13,111 (43.8) | 14,364 (44.9) |
| Medicaid | 17,442 (28.2) | 8,728 (29.2) | 8,714 (27.3) |
| Private | 15,476 (25.0) | 7,438 (24.9) | 8,038 (25.1) |
| Self-pay | 1,097 (1.8) | 431 (1.4) | 666 (2.1) |
| Other | 390 (0.6) | 204 (0.7) | 186 (0.6) |
| Unknown | 3,116 | 1,520 | 1,596 |
| Discharge disposition, No. (%) |  |  |  |
| Home | 44,511 (68.5) | 21,176 (67.4) | 23,335 (69.5) |
| Home with home health | 11,267 (17.3) | 5,739 (18.3) | 5,528 (16.5) |
| Skilled nursing facility/rehab | 5,572 (8.6) | 2,760 (8.8) | 2,812 (8.4) |
| Hospice/expired | 1,932 (3.0) | 928 (3.0) | 1,004 (3.0) |
| Against medical advice | 797 (1.2) | 412 (1.3) | 385 (1.1) |
| Long-term acute care | 336 (0.5) | 165 (0.5) | 171 (0.5) |
| Other transfer | 260 (0.4) | 98 (0.3) | 162 (0.5) |
| Psychiatric facility | 197 (0.3) | 104 (0.3) | 93 (0.3) |
| Urine drug screen, No. (%) | 7,146 (11.0) | 3,544 (11.3) | 3,602 (10.7) |
| Amphetamine/methamphetamine | 169 (2.4) | 84 (2.4) | 85 (2.4) |
| Barbiturates | 232 (3.2) | 121 (3.4) | 111 (3.1) |
| Benzodiazepine | 1,106 (15.5) | 534 (15.1) | 572 (15.9) |
| Cocaine/metabolite | 886 (12.4) | 416 (11.7) | 470 (13.0) |
| Ecstasy | 28 (0.4) | 14 (0.4) | 14 (0.4) |
| Marijuana/tetrahydrocannabinol | 1,450 (20.3) | 716 (20.2) | 734 (20.4) |
| Methadone | 283 (4.0) | 152 (4.3) | 131 (3.6) |
| Opiates | 1,321 (18.5) | 681 (19.2) | 640 (17.8) |
| Oxycodone | 114 (1.6) | 43 (1.2) | 71 (2.0) |
| Phencyclidine | 71 (1.0) | 37 (1.0) | 34 (0.9) |
| Blood alcohol concentration testing, No. (%) | 1,870 (2.9) | 959 (3.1) | 911 (2.7) |
| Blood alcohol concentration level, No. (%) |  |  |  |
| 0% | 1,513 (81.0) | 748 (78.1) | 765 (84.0) |
| ≤0.10% | 93 (5.0) | 49 (5.1) | 44 (4.8) |
| 0.10-0.20% | 73 (3.9) | 45 (4.7) | 28 (3.1) |
| >0.20% | 190 (10.2) | 116 (12.1) | 74 (8.1) |

### SMART-AI Screen-Positive Classification

During the SMART-AI period, 1,476 of 33,564 hospitalizations(4.4%) were flagged by the algorithm as screen-positive for substance misuse based on clinical documentation from the first 24 hours of admission.. Of all SUIT consultations ordered during the post-period, 71.4% (n=801) had screened positive by SMART-AI. Among the 33,564 hospitalizations during the SMART-AI period, the algorithm flagged 940 for opioid misuse (2.8%), 581 for alcohol misuse (1.7%), and 15 for nonopioid drug misuse (0.04%) (**Table 2**).

**Table 2.** Screening and consult outcomes.

| **Characteristic** | **Overall**<br>**(N = 64,996)** | **Usual Care**<br>**(N = 31,432)** | **SMART-AI**<br>**(N = 33,564)** |
| --- | --- | --- | --- |
| Consult ordered, No. (%) | 2,568 (4.0) | 1,295 (4.1) | 1,273 (3.8) |
| Addiction medicine note, No. (%) | 2,347 (3.6) | 1,196 (3.8) | 1,151 (3.4) |
| Consult completed, No. (%)<sup>1</sup> | 2,299 (3.5) | 1,178 (3.7) | 1,121 (3.3) |
| Hours to consult, median (IQR) | 15 (7-29) | 14 (5-27) | 16 (8-31) |
| Social work brief intervention/motivational interviewing, No. (%) | 523 (0.8) | 246 (0.8) | 277 (0.8) |
| Naloxone, No. (%)<sup>2</sup> | 444 (0.7) | 282 (0.9) | 162 (0.5) |
| Medication-assisted treatment, No. (%) | 1,390 (2.1) | 726 (2.3) | 664 (2.0) |
| Alcohol use disorder | 504 (36.3) | 277 (38.2) | 227 (34.2) |
| Opioid use disorder | 894 (64.3) | 443 (61.0) | 451 (67.9) |
| SMART-AI positives, No. (%) |  |  |  |
| Opioid | -- | -- | 940 (2.8) |
| Alcohol | -- | -- | 581 (1.7) |
| Nonopioid | -- | -- | 15 (0.0) |
| Any substance use | -- | -- | 1,476 (4.4) |
| Composite, No. (%)<sup>3</sup> | 2,333 (3.6) | 1,189 (3.8) | 1,144 (3.4) |
| Readmission among those who received the composite outcome, No. (%)<sup>4</sup> | 900 (39.2) | 485 (41.3) | 415 (37.0) |

<sup>1</sup>Consult ordered and consult or care plan note type present.
<sup>2</sup>Prescription, free kit, or take-home order.
<sup>3</sup>Medication-assisted treatment, social work brief intervention/motivational interviewing, naloxone dispensing, or consult completed.
<sup>4</sup>Those discharged to hospice or died in hospital excluded

### AI-Assisted Workflow

Over the post-implementation period, a total of 624 SMART-AI screen positives were reviewed by SUIT for potential follow-up as part of the study (**Figure 1**). SUIT reviewed positives that were still hospitalized and missing a consult order at the time they reviewed the daily list. 55 of these were noted as false positives after chart review. For 271 of those that SUIT agreed were appropriate for consult, an Epic Secure Chat message was sent to the primary team, and 96 subsequently completed consults (**Figure 1**).

### Primary Outcome

At least one component of the composite intervention was received in 1,189 of 31,432 hospitalizations (3.8%) during usual care and 1,144 of 33,564 (3.4%) during the SMART-AI period (absolute difference, −0.4 percentage points; 95% CI, −0.7 to −0.1; P = 0.20) (**Figure 2**). The lower bound of the 95% CI (−0.7) crossed the prespecified noninferiority margin of −0.5. Naloxone was dispensed at discharge to 0.9% vs 0.5%, MAT was initiated or adjusted in 2.3% versus 2.0%, and brief intervention or motivational interviewing was documented in 0.8% for both periods (**Table 2**). Among hospitalizations receiving at least one component of the composite intervention (N = 2,333), patients in the SMART-AI period were older (mean, 50.4 vs 47.9 years) and had a higher proportion of Black individuals (46.0% vs 40.9%) compared with the manual period. Inpatient medication exposures also differed, with greater opiate administration in the SMART-AI cohort (36.5% vs 31.7%) and lower benzodiazepine use (14.6% vs 23.9%), along with reduced exposure to dexmedetomidine (5.3% vs 7.3%) and propofol (4.4% vs 6.2%) (**eTable 1**).

**Figure 2:**
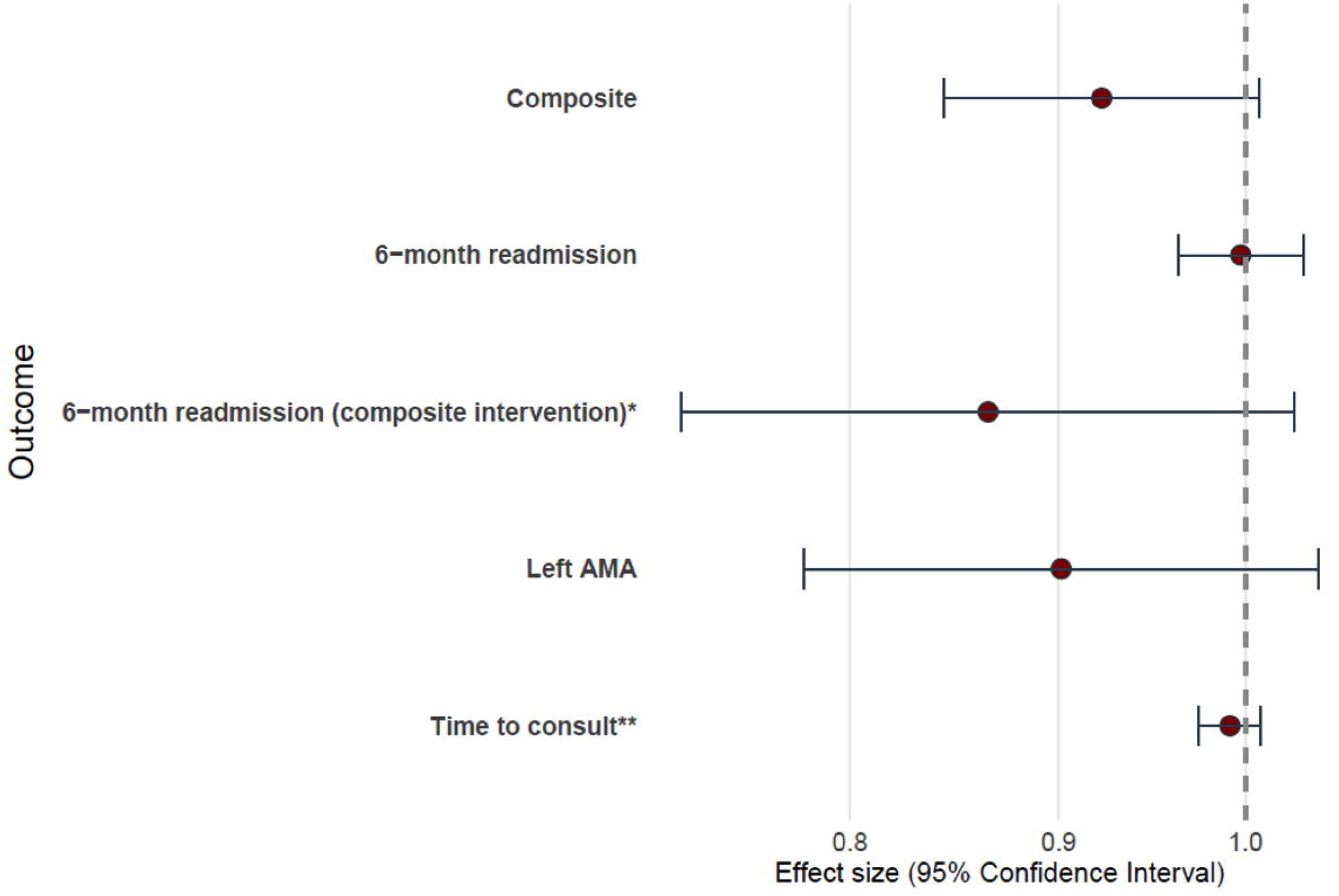
Adjusted Effects of SMART-AI Implementation on Clinical and Utilization Outcomes. The forest plot displays adjusted effect estimates (points) with 95% confidence intervals (horizontal bars) comparing the post-implementation period with AI-assisted screening (SMART-AI) to the pre-implementation period with manual screening. Outcomes include the composite clinical intervention (receipt of at least one of the following after SUIT involvement: naloxone dispensing; initiation or adjustment of medication for alcohol or opioid use disorder; brief motivational intervention; or completed addiction-medicine consultation), 6-month readmission, 6-month readmission among those who received a component of the composite intervention*, time to consult**, and discharge against medical advice (left AMA). Estimates are derived from multivariable models adjusting for age, sex, race and ethnicity, insurance status, and hospital unit; error bars represent 95% CIs. The vertical dashed line indicates the null value (ratio = 1.0); values less than 1 favor SMART-AI, while values greater than 1 favor manual screening. AMA stands for “against medical advice.”* Time-to-consult is modeled as a rate/time ratio; values less than 1 indicate a shorter time to consult during the SMART-AI period.

### Secondary Outcomes

The median time from admission to consultation completion did not differ significantly between periods (14 hours [IQR, 5-27] vs 16 hours [IQR, 8-31]; rate ratio: 0.99, 95% CI: 0.97-1.01, p = 0.33). Discharge against medical advice occurred in 1.3% (412/31,432) during usual care versus 1.1% (385/33,564) during SMART-AI (odds ratio: 0.99, 95% CI: 0.78-1.04, p=0.16). Adjusted analyses showed no difference in all-cause hospital-wide 6-month readmission between periods (odds ratio: 1.00, 95% CI: 0.96–1.03; P=0.89). Among patients who received a SUIT intervention (n = 2,296), 6-month readmission occurred in 41.3% (485/1,175) during usual care versus 37.0% (415/1,121) during SMART-AI (odds ratio: 0.86, 95% CI: 0.73-1.03, p=0.10).

### Cost-Effectiveness Analysis Program Setup and Maintenance

The initial planning and implementation of the manual screening program involved fixed costs of $30,807. These included training unit social workers (n = 23) in brief intervention and motivational interviewing, along with monthly maintenance costs of $457 (**Table 3; eTable 2 and 3**). For the SMART-AI program, fixed costs were $10,429; monthly expenses to maintain the AI model and review potential errors amounted to $3,230, while monthly variable costs to operate the AI model within the existing workflow added $980.54 for a total monthly cost of $4,210 (eTable 4).

**Table 3.**
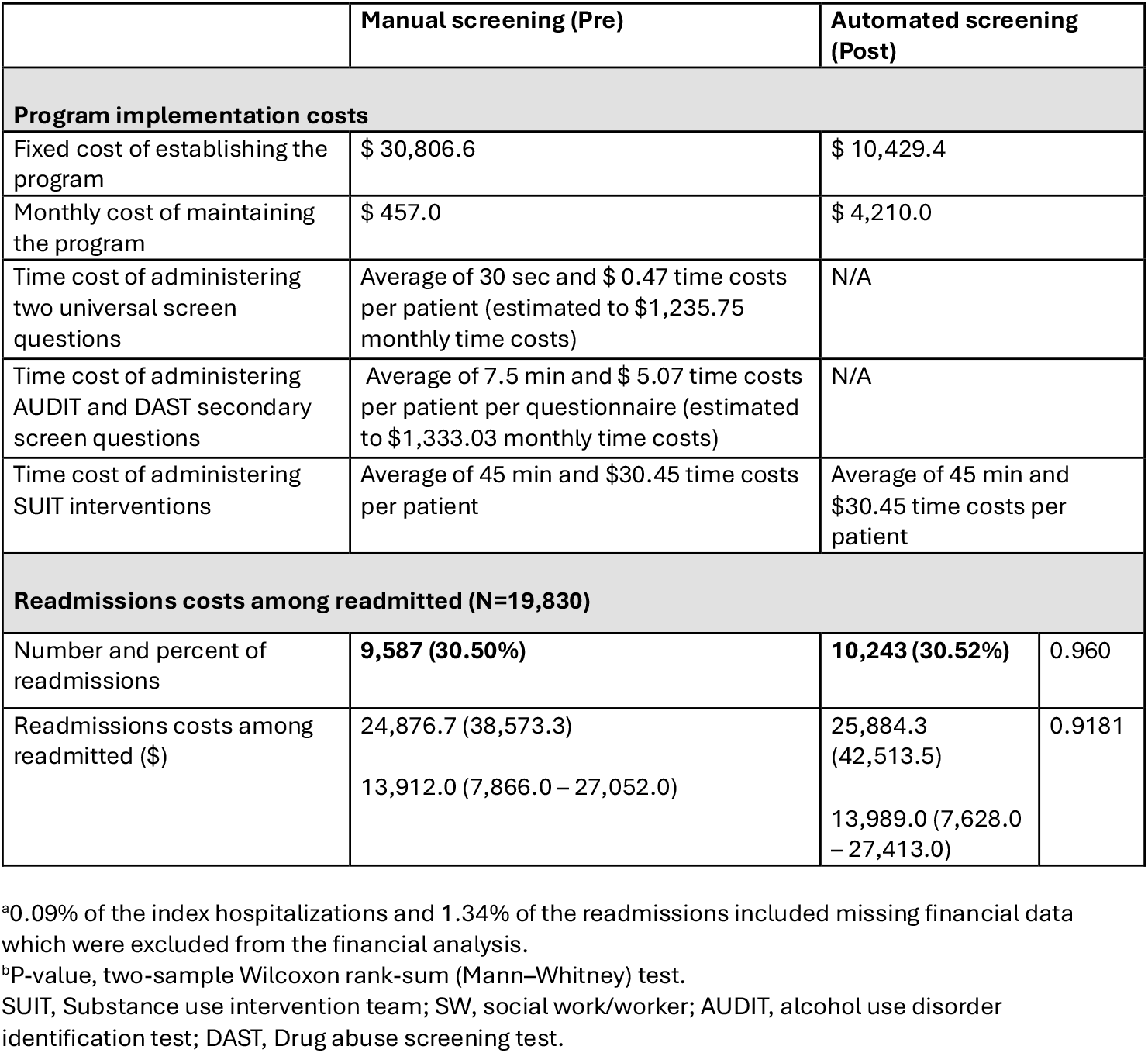
Program implementation, hospitalization, and readmission costs (2024 U.S. $)

### Task-Level Time and Cost

Administering the manual screening program with the brief, universal screening questions by nurses took between 15 and 45 seconds, costing $0.23 to $0.70 per screening (average, 30 seconds; $0.47). Administering the full screening instrument by unit social workers took 5 to 10 minutes per instrument (alcohol and opioids/drugs), costing $3.38 to $6.77 per patient per questionnaire (average, 7.5 minutes; $5.07) (**eTable 5**). Brief intervention and motivational interviewing delivered by the SUIT team social workers required 30 to 60 minutes, costing $20.30 to $40.60 per patient (average, 45 minutes; $30.45) (**eTable 6**).

### Hospitalization Costs

Median costs for the index hospitalization were $15,229 (IQR, $8,665-$28,908) during usual care and $14,752 (IQR, $8,144-$28,052) during the SMART-AI period. Median costs for readmission hospitalizations were $13,912 (IQR, $7,866-$27,052) during usual care and $13,989 (IQR, $7,628-$27,413) during the SMART-AI period.

### Incremental Cost-Effectiveness

Over the course of one year of implementation, the total annual program costs for automated screening were $6,167 less than those for manual screening. The ICER for the composite outcome was −$77.51 per unit change during automation (95% CI, −$133.98 to $896.18), which was −$49.55 (95% CI, −$154.38 to $554.08) when fewer universal screenings occurred during usual care (**eTable 7**). Automated screening was associated with total program cost reductions compared to manual screenings while maintaining similar readmission outcomes. Cost savings were estimated to be greater when the analysis focused on patients who received addiction medicine services, since larger readmission reductions were observed in this subgroup (**Table 2**).

## Discussion

In this first large-scale evaluation of automated screening for multiple types of substance misuse, automation maintained readmission rates and reduced program costs, though the prespecified noninferiority criterion was not met. While 79% of U.S. hospitals report offering substance misuse screening services, this reported availability does not translate to consistent implementation^28^. Barriers, including financial strain, inadequate personnel, stigma, lack of expertise, and time constraints, result in substantial variability, with hospital-level screening rates ranging from as low as 12% to as high as 65%.^28– 30^ This automated screening approach, requiring only secure EHR messaging, may enable scalable substance misuse screening across U.S. hospitals. Although the prespecified noninferiority criterion was not met, AI-based automation achieved similar clinical outcomes to manual screening at lower cost, suggesting this approach could replace labor-intensive workflows where hospitals struggle with consistent implementation or establish screening programs where none currently exist.

This wide variability in screening rates reflects substantial hospital-level differences in implementation success ^28,29^. Organizational factors, including staffing availability, competing clinical priorities, and clinician discomfort with screening, contribute to inconsistent adoption^30,31^. Even at hospitals with screening programs, approximately half of admitted patients may be screened due to staffing limitations, and patients rarely disclose problematic substance use to providers who infrequently use validated screening tools ^32^. Time and workflow constraints, coupled with stigma and lack of clinician knowledge, further limit the adoption of evidence-based counseling following positive screens^33^. Automated screening processes review all admission notes overnight and flag high-risk patients for expert review, eliminating the need for nurse or social worker time. This enables universal screening, independent of patient disclosure or clinician comfort, while addressing the operational barriers that prevent the consistent implementation of manual screening programs.

Addiction medicine consultations reduce morbidity and readmissions in hospitalized patients with substance use disorders, but few studies have evaluated screening tools based on AI^34–36^. A prior automated screening study for opioid misuse alone met its non-inferiority criterion and reduced 30-day readmissions, but required EHR vendor collaboration, custom alert logic, and real-time processing, triggering alerts a median of 10 times per hospitalization^14^. In contrast, our multilabel model screened for alcohol, opioid, and non-opioid substances simultaneously, reflecting polysubstance use patterns common in hospitalized patients, while requiring only basic EHR messaging through overnight batch processing with a single daily secure chat notification to the SUIT team. This dual human-in-the-loop architecture may have reduced intervention rates while providing clinical oversight and avoiding alert fatigue^37^. Implementation architecture—not just model performance—determines clinical impact, with trade-offs between maximizing reach through direct alerts and optimizing appropriateness through expert review.

Automated screening was cost-saving, reducing annual program costs by over $6,000 and achieving a favorable incremental cost-effectiveness ratio—lowering costs while maintaining similar clinical outcomes, with even greater savings among patients who received addiction services. Computational resource costs are decreasing over time, further supporting the economic viability of our approach. The minimal integration requirements of our approach, relying only on basic EHR messaging without vendor collaboration or custom development, make it more transferable to community hospitals and health systems with limited informatics resources. Depending on technical capacity and institutional priorities, health systems can choose full EHR integration with best practice advisories or a lighter implementation using secure chat.

## Limitations

This single-center study without concurrent controls is susceptible to temporal confounding, although the 12-month baseline and covariate adjustment mitigate this risk. Because automated screening and SUIT selective outreach were implemented together, the independent effect of each component cannot be isolated. The dual human-in-the-loop workflow may have contributed to lower intervention rates, and it remains unclear whether reduced delivery reflects appropriate triage or missed opportunities, given that the lower bound of the noninferiority margin was not met. The composite outcome may also obscure variation across its components. In addition, outcomes may reflect the SUIT team’s clinical expertise more than the screening tool itself, limiting generalizability to settings without comparable resources. The consensus-based noninferiority margin was not empirically derived, and observed cost reductions may partly reflect broader institutional cost-containment efforts. Despite these limitations, outcomes remained similar across periods, program costs were lower with automation, and the findings demonstrate that large-scale automated screening is feasible.

## Conclusions

In a study of 64,996 hospitalizations, AI-assisted automated screening for substance misuse did not meet non-inferiority criteria but achieved secondary outcomes comparable to those of manual screening at a reduced cost. The minimal integration approach, requiring only secure EHR messaging, offers a feasible and economically viable pathway for hospitals where manual screening programs are absent or unsustainable. Given the substantial gap between reported program availability and actual screening implementation, this lower-barrier approach may enable broader and more consistent screening nationally. Future studies should compare implementation strategies across diverse settings, integrating human factors engineering, AI-augmented workflow designs, and behavioral economics to identify the most effective approaches.

## Data Availability

All data produced in the present study are available upon reasonable request to the authors

## Author Contributions

Concept and design: Rojas, Joyce, Afshar, Dligach, Churpek. Acquisition, analysis, or interpretation of data: Rojas, Joyce, Markossian, Chaudhari, McClintic, Castro, Fairgrieve, Dligach, Oguss, Churpek, Nikolaides, Afshar. Drafting of the manuscript: Rojas, Joyce, Markossian, Afshar. Critical revision of the manuscript for important intellectual content: All authors. Statistical analysis: Joyce, Markossian, Afshar. Administrative, technical, or material support: Chaudhari, McClintic, Castro, Fairgrieve, Oguss. Supervision: Afshar, Churpek. Data access, responsibility, and integrity: Rojas, Joyce, Markossian, Afshar had full access to all the data in the study and take responsibility for the integrity of the data and the accuracy of the data analysis. Final approval of the manuscript: All authors.

