## Supplement for "Clinical Implementation of an AI Algorithm for Substance Misuse Screening in Hospitalized Adults"

### SUPPLEMENTAL DATA

**Table E1. Baseline Characteristics of Patients in Composite Outcome Analysis**

| Characteristic | Overall (N = 2,333) | Usual Care (N = 1,189) | SMART-AI (N = 1,144) |
|---|---|---|---|
| Age, mean (SD) | 49.2 (13.9) | 47.9 (13.6) | 50.4 (14.1) |
| Sex, No. (%) | | | |
| Female | 736 (31.5) | 377 (31.7) | 359 (31.4) |
| Male | 1,597 (68.5) | 812 (68.3) | 785 (68.6) |
| Race/ethnicity, No. (%) | | | |
| Asian | 6 (0.3) | 3 (0.3) | 3 (0.3) |
| Black | 998 (43.4) | 479 (40.9) | 519 (46.0) |
| Hispanic | 436 (19.0) | 219 (18.7) | 217 (19.2) |
| Other | 118 (5.1) | 55 (4.7) | 63 (5.6) |
| White | 742 (32.3) | 415 (35.4) | 327 (29.0) |
| Patient class, No. (%) | | | |
| Inpatient | 1,768 (75.8) | 890 (74.9) | 878 (76.7) |
| Observation | 565 (24.2) | 299 (25.1) | 266 (23.3) |
| Admission type, No. (%) | | | |
| Elective | 240 (10.3) | 109 (9.2) | 131 (11.5) |
| Emergency | 2,093 (89.7) | 1,080 (90.8) | 1,013 (88.5) |
| Admission origin, No. (%) | | | |
| Direct admission | 52 (2.2) | 31 (2.6) | 21 (1.8) |
| ED admission | 1,946 (83.4) | 1,011 (85.0) | 935 (81.7) |
| OR admission | 82 (3.5) | 35 (2.9) | 47 (4.1) |
| Transfer | 253 (10.8) | 112 (9.4) | 141 (12.3) |
| Insurance, No. (%) | | | |
| Medicare | 406 (17.7) | 183 (15.6) | 223 (19.9) |
| Medicaid | 1,561 (68.1) | 826 (70.4) | 735 (65.7) |
| Private | 209 (9.1) | 115 (9.8) | 94 (8.4) |
| Self-pay | 109 (4.8) | 46 (3.9) | 63 (5.6) |
| Other | 6 (0.3) | 3 (0.3) | 3 (0.3) |
| Length of stay, days, mean (SD) | 6.1 (6.7) | 6.3 (7.0) | 5.9 (6.4) |
| Elixhauser comorbidity score, mean (SD) | 1.9 (6.2) | 1.9 (6.2) | 2.0 (6.2) |

| **Table E2**. Marginal total costs, effectiveness and incremental cost effectiveness ratio (ICER) of automated (Post) versus manual (Pre) screening over one year | | | | | | | |
|---|---|---|---|---|---|---|---|
| **Scenario** | **Costs included** | **Outcome (adjusted)** | **Marginal probability of outcome (Post versus Pre) (adjusted)** | **ICER Numerator: Delta Costs (estimated difference in total costs) (Post – Pre)** | **ICER Denominator: Delta Outcome (estimated relative difference in outcome) (Post – Pre)** | **ICER (CI, Confidence Interval)** | **ICER quadrant and interpretation** |
| 1 | • Program fixed and monthly maintenance costs (Pre and Post periods)<br>• Manual screening time costs (Pre period): for 100% universal screening during all index hospitalizations and 10% secondary screening either with AUDIT or DAST | **Composite Outcome (receipt of addiction medicine services)** | -0.0025 | -6,166.71 | -79.56 | $77.51<br>(CI, -133.98 – 896.18) decrease per unit change in outcome | The intervention is less costly |
| 2 | • Program fixed and monthly maintenance costs (Pre and Post periods)<br>• Manual screening time costs (Pre period): for 85% universal screening during all index hospitalizations and 10% secondary screening either with AUDIT or DAST | **Composite Outcome (receipt of addiction medicine services)** | -0.0025 | -3,942.35 | -79.56 | $49.55<br>(CI, -154.38 – 554.08) decrease per unit change in outcome | The intervention is less costly |

| | | | | | | | |
|---|---|---|---|---|---|---|---|
| 3 | • Programs fixed and monthly maintenance costs (Pre and Post periods)<br>• Manual screening time costs (Pre period): for 100% universal screening during all index hospitalizations and 10% secondary screening either with AUDIT or DAST<br>• Time cost of administering addiction medicine services by SUIT team (Pre and Post periods) | **Readmission** | -0.00048 | -8,589.21 | -15.04 | N/A | Cost saving, the intervention is dominant to the control because the intervention is less costly, and it maintains and slightly reduces (albeit not statistically significant) readmissions |
| **Analysis among hospitalized patients who received addiction medicine services (n=2,296)** | | | | | | | |

| | | | | | | | |
|---|---|---|---|---|---|---|---|
| 4 | • Programs fixed and monthly maintenance costs (Pre and Post periods)<br>• Manual screening time costs (Pre period): for 100% universal screening during all index hospitalizations and 10% secondary screening either with AUDIT or DAST<br>• Time cost of administering addiction medicine services by SUIT team (Pre and Post periods) | **Readmission** | -0.032004 | -8,589.21 | -36.63 | N/A | Cost saving, the intervention is dominant to the control because the intervention is less costly, and it maintains and slightly reduces (albeit not statistically significant) readmissions |
| ICER, incremental cost effectiveness ratio; CI, Confidence Interval; s.d., standard deviation; AUDIT, alcohol use disorder identification test; DAST, Drug abuse screening test; SUIT, Substance use intervention team. | | | | | | | |
| Average eligible admissions per month (s.d.) = 2,629.26 (349.9) | | | | | | | |
| Average eligible admissions per month among who received addiction medicine services (s.d.) = 95.25 (16.08) | | | | | | | |

**Table E3**. Roles of personnel involved in substance misuse screening and interventions during pre (manual) and post (automated) substance misuse screening in a large urban academic medical center (2024 U.S. $).

| Personnel | Role | Base Salary[a] (based on regional averages)/Base Salary + Fringe (30%) ($) | Rate per hour including fringe[b] ($) | Program phase |
|---|---|---|---|---|
| SUIT Medicaldirector | Lead the planning and implementation of the program. [Emergency, addiction and toxicology medicine physician] | $240,000 / $312,000 | $150.0 | Pre and Post |
| SUIT SW lead | Training and supporting SWs | $80,000 / $104,000 | $50.0 | Pre and post |
| SUIT SWs | Receive training, and administer SBIRT and other SW interventions | $65,000 / $84,500 | $40.6 | Pre and Post |
| Epic EHR application analyst | Built flowsheets and programs in Epic EHR | $90,000 / $117,000 | $56.2 | Pre |
| Epic EHR data scientist | Built advanced analytics for AI tool implementation in Epic EHR | $90,000 / $117,000 | $56.2 | Post |
| Nursing director | Planning collaboration | $200,000/ $260,000 | $125.0 | Pre |
| Case management-SW director | Planning collaboration | $125,000 / $162,500 | $77.9 | Pre |
| Nurse (ED or inpatient) | Administer 2 questions manual screen | $90,000 / $117,000 | $56.2 | Pre |
| SW (floor or SUIT team) | Administer secondary screen | $65,000 / $84,500 | $40.6 | Pre |
| Chief Medical Informatics Officer | Planning collaboration | $240,000 / $312,000 | $150.0 | Post |
| Medical informatics physician champion | Lead the planning and implementation of the automated AI tool. [Internal medicine physician] | $240,000 / $312,000 | $150.0 | Post |
| Automation program coordinator | Help establish and implement workflow for AI screening in clinical practice. | $60,000 / $78,000 | $37.5 | Post |

a. Median salary in large Midwestern metropolitan area in 2024, available from salary.com and reviewed and validated by practitioners on the study team.
b. Based on 40 hours workweek that is equivalent to 2,080 hours/year.

EHR, electronic health record; SUIT, Substance use intervention team; SW, social work/worker; AUDIT, alcohol use disorder identification test; DAST, Drug abuse screening test.

**Table E4**. List of activities, definitions, and costs associated with establishing the manual substance misuse screening (fixed costs) (2024 U.S. $)

| Activities | Definitions and functions | Role | Cost driver | Cost per activity |
|---|---|---|---|---|
| Plan the implementation of the substance misuse universal screening in all inpatient units | Secure buy-in from all involved departments (SUIT, case management - SW, nursing) and planning the process. | SUIT medicaldirector, nursing director, Case management-SW director, and SUIT SW lead. | Bi-weekly 1-hour long meetings over 3-months, for a total of 6 hours per attendee. | 6 x $150.0 + 6 x $125.0 + 6 x $77.96 + 6 x $50.0 = $2,417.8 |
| Building epic flowsheet and necessary adjustments | Build customized Epic flowsheet for each staff role involved in manual screening and implement necessary improvements over time | Epic application analyst | 50 hours | 50 x $56.2 = $2,810.0 |
| Deliver and receive training for AUDIT and DAST screening | In-person training of all inpatient unit SWs, training administered by SUIT SW lead | All inpatient case manager-social workers (23 FTE) + trainer (1 FTE) | 2-hours long training | 2 x 23 x $40.6 + 2 x $50.0 = $1,967.6 |
| Training for brief intervention and motivational Interviewing (BI/MI) | In-person training of all unit SWs, training administered by SUIT SW lead | All inpatient case manager-social workers (23 FTE) + trainer (1 FTE) | 3-days long training; 24 hours | 24 x 23 x $40.6 + 24 x $50.0 = $23,611.2 |
| | | | **Total Fixed costs** | **$30,806.6** |

SUIT, Substance use intervention team; SW, social work/worker; AUDIT, alcohol use disorder identification test; DAST, Drug abuse screening test.

**TableE5**. List of activities, definitions and monthly costs associated with maintaining the manual substance misuse screening (fixed monthly costs) (2024 U.S. $)

| **Activities** | **Definitions and functions** | **Personnel and FTEs** | **Cost drivers** | **Cost per activity per month** |
|---|---|---|---|---|
| Program monitoring and evaluation | Monthly meetings between SUIT medical director, SUIT SW lead and case management-SW director | SUIT medical director, SUIT SW lead and social work director | Monthly, 1-hour long meetings | $150.0 + $50.0 + $77.9 = $277.9 |
| Bi-yearly refresher training of SWs | Refresher training for AUDIT/ DAST/ and BI/MI and feedback about program | All inpatient case manager- social workers (23 FTE) + SUIT SW trainer (1 FTE) | 1-hour long refresher training every 6 months | 2 x 23 x $40.6 + 2 x $50.0 = $1,967.6/ 12 = $164.0 |
| New SW hire onboarding | Training of AUDIT/ DAST/ and BI/MI | New hire social worker (2 FTE) + SUIT SW trainer (1 FTE) | 1-hour long onboarding | 1 x 2 x $40.6 + 2 x $50.0 = $181.2 / 12 = $15.1 |
| | | | **Total Fixed Monthly costs** | **$457.0** |

SUIT, Substance use intervention team; SW, social work/worker; AUDIT, alcohol use disorder identification test; DAST, Drug abuse screening test; BI/MI, brief intervention and motivational Interviewing.

**Table E6**. List of activities, definitions, and costs associated with establishing the automated substance misuse screening (fixed costs) (2024 U.S. $)

| Activities | Definitions and functions | Role | Cost driver | Cost per activity |
|---|---|---|---|---|
| Design a clinical workflow to incorporate feedback from the smart AI tool to clinical practice. | Discussions to establish processes to implement feedback from the smart AI tool in clinical practice, and silent testing of AI screening tool. | SUIT medical director, medical informatics physician champion, AI coordinator and SUIT SWs (2 FTEs) | Two 1-hour long meetings over 1-month, for a total of 2 hours per attendee, including email communications. | 2 x $150.0 + 2 x $150.0 + 2 x $37.5 + 2 x 2 x $40.6 = $837.4 |
| Discuss options to integrate AI tool within Epic and communicate AI output with clinical team. | Discuss potential implementation of AI tool within the medical center and get the buy-in and agreement from hospital IT department chief. | Chief medical informatics officer and medical informatics physician champion | 1-hour long meetings, 2 times | 2 x $150.0 + 2x $150.0 = $600.0 |
| Build the smart AI tool pipeline in Epic using Azure | Using Azure, build the AI tool pipeline in Epic and deploy by generating daily automated email that includes information of all hospital admissions classified as inpatient or observation (from the previous 48 hours) with AI algorithm score along flag of patients with possible substance misuse. | Epic EHR Data Scientist (1 FTE) | 160 hours | 160 x $56.2 = $8,992.0 |
| | | | **Total Fixed costs** | **$ 10,429.4** |

AI, artificial intelligence; SUIT, Substance use intervention team; SW, social work/worker.

**Table E7**. List of activities, definitions and monthly costs associated with maintaining and administering the automated substance misuse screening[a] (fixed and variable monthly costs) (2024 U.S. $)

| Activities | Definitions and functions | Personnel and FTEs | Cost drivers | Cost per activity per month |
|---|---|---|---|---|
| AI tool maintenance | Maintaining the automated AI tool and reviewing possible errors | Medical informatics physician champion, AI program coordinator and Epic EHR data scientist, it also includes additional work by data scientist to implement the discussed changes in AI tool pipeline. | Monthly, 15 min long meeting and/or communication between medical informatics physician champion and AI program coordinator, and data scientist (more extensive communication during months 1 & 2 of implementation, and less frequent during months 3-12) + additional monthly work by the data scientist to implement discussed changes in AI tool pipeline (months 1 & 2 are 8 hours a month and then 2 hours a month 3-12). | 0.25 hours x ($150.0 + $56.2 + $37.5) + 3 hours x $56.2 = $229.53 |
| AI tool Pipeline Computing Resources (Microsoft application Azure) | Storage and Processing, monthly fees allocated by the health system to the cost unit. | | Azure Resource Group Monthly Cost | $3,000 |
| | | | **Fixed Monthly costs** | **$ 3,229.5** |
| Identify and communicate potential missed patients with SUIT team. | Receives the automated email, conduct initial chart reviews of patients who are tagged by AI, and then sends relevant patient information to SUIT SWs in a secure message. | AI program coordinator | It takes on average 30 minutes each day to complete this work. [There are about 1 to 15 patients in the excel sheet from the automated message generated by the AI for an average of 8 patients each day, and about 3-5 min to review each patient chart]. | 30.5 days x 0.5 hours x $37.5 = $571.9 |

| | | | | |
|---|---|---|---|---|
| SUIT team review of missed patients and place SUIT consults. | Review the charts of missed patients, and if appropriate, send an EHR message to the primary care team about possible SUIT consult. | SUIT SWs | It takes on average 20 minutes each day to complete this work. [There are on average 4-5 missed patients every day, and it takes SW about 4 min per patient to review the chart and place HER secure message to primary care team] | 30.5 days x 0.33 hours x $40.6 = $408.64 |
| | | | **Total Monthly costs** | **$ 4,210.0** |

a. Calculated from two SUIT SWs and AI program coordinator interviews.

AI, artificial intelligence; SUIT, Substance use intervention team; SW, social work/worker.

**TableE8**. Variable cost drivers of administering manual substance misuse screenings (2024 U.S. $)

| Activities | Personnel | Cost drivers |
|---|---|---|
| Two questions universal screen (embedded in the admissions database) | Nurse | 15 – 45 sec for a time cost of $0.23 - $0.70 per screening (average 30 sec and time cost of $0.47)[b] |
| Secondary screen with AUDIT and DAST | Social Work (SW) (floor) | 5-10 min per questionnaire for a time cost of $3.38 - $6.77 per patient per questionnaire (average 7.5 min and $5.07 per patient per questionnaire)[c] |

a. These will be added, as appropriate, to hospitalization total costs (since these are non-billable services).
b. Calculated from 3 intensive care unit (ICU) and 3 floor staff nurse interviews.
c. Calculated from 5 SW interviews.

**TableE9**. Variable cost drivers of administering SUIT interventions (2024 U.S. $)

| Activities | Personnel | Cost drivers |
|---|---|---|
| SUIT Interventions | SW (SUIT) | Interventions by SUIT SWs takes about 30-60 min for a cost of $20.3 – $40.6 per patient (average 45 min and $30.45 per patient).<br><br>Breakdown of SUIT interventions include:<br>1 - Motivational interviewing: ~30 min<br>2 - Planning MAT follow-up: ~15 min<br>3 – Discussion of SUD treatment options: ~15-30 min<br>4 – Direct transfer to SUD treatment program: ~30 – 45 min<br>5 - Care coordination with post-acute care facility: ~ 5-10 min<br>6 - Other disposition planning (transportation, other referrals, etc.): ~5-10 min<br><br>Most frequently the SW would do 1, 2 & 3. |

a. These will be added, as appropriate, to hospitalization total costs (since these are non-billable services).
b. Calculated from 3 SUIT SW interviews.

### Cost Interview Guides

***GUIDE 1:*** *Interview guide for SUIT personnel regarding costs associated with establishing and maintaining universal manual substance misuse screening (SUIT medical director, SUIT social worker lead, SUIT social worker) – not all questions may apply to all interviewees. .*

**Section 1**: SUIT Program timeline

Q1: When was the start date of the SUIT program at [health system name], can you explain briefly how did the program evolve over-time, for example, the stages of manual screening?

Q2: Can you describe briefly what does it take to plan and implement the SUIT program?
Did you hire any personnel dedicated for the program, and if so, what's their role, credentials and contribution to the program?

Q3: Can you describe briefly all the personnel involved in the SUIT, both for screening and treatment, credentials and roles?
Program director?
Coordinator?
Social Worker?
Nurses at the Units, type?
Doctors?

**Section 2**: SUIT program personnel training

Q4: Can you describe the initial hospital-wide training process to start the SUIT program for screening and treatment, if any?
What's in the training material?
Who created the training material? How long it took to create the training material?
Who administered the trainings, how frequently, and who participated?
Did you hire any consultants or send personnel to attend trainings?

Q5: Do you conduct any ongoing trainings? Can you describe what's in the trainings, who delivers them and who attends them?

**Section 3**: SUIT program screening

Q6: Who conducts the universal screenings (U.S.), floor nurses or SUIT social workers? Where do you document about the results of the U.S.?

Can you estimate the percent of universal screens administered by SW?

Can you estimate the percent of U.S. administered by floor nurses?

Anyone else administers U.S., and if so, who and what % of the U.S.?

Q7: Who conducts the secondary screen (AUDIT/DAST)? Where do you document about the results of the secondary screen?

Q8: Who conducts the Brief Intervention/MI (mild/moderate condition vs. severe condition), Inpatient SUIT Consults, Inpatient MATs, and outpatient SUIT consults?

Q9: Do you charge for Brief Intervention/MI, SUIT Consults, Inpatient MATs, and outpatient SUIT consults?

Can you describe <u>all</u> the CPT (and/or HCPCS) codes associated with these services?
Can you describe <u>all</u> the ICD9 and 10 (and/or HCPCS) codes associated with these services?

***GUIDE 2:*** *Interview guide regarding costs associated with establishing and maintaining automated substance misuse screening (AI program coordinator, SUIT medical director, SUIT social workers) – not all questions may apply to all* *interviewees.*

**Section 1:** Designing clinical workflow to incorporate feedback from the smart AI tool to clinical practice

Q1: Who are the personnel involved in building the smart AI tool pipeline in Epic, credentials and roles?

Program director, Informatics team members, physician champions, nursing, anyone else?
How frequently and how long did you meet?
How many hours per week the informatics team spent building the AI tool pipeline in Epic?
What is the platform you used to build the AI tool in Epic?
Any need for IT work to maintain the AI tool pipeline?

Q2: Who are the personnel involved in planning and implementing the integration of AI tool and output within clinical workflow, credentials and roles?

Program director, Informatics team members, physician champions, nursing, anyone else?
How frequently and how long did you meet?
How many hours per week the informatics team spent building and updating communication pathways between output from the AI tool pipeline with clinical team?

**Section 2**: Initial and ongoing training for adoption and use of AI tool pipeline

Q3: Can you describe process to inform hospital staff [who - physicians, nurses, SW, others] in adopting and using the smart AI tool in practice?
Who is using output from the AI tool?

How did you collect feedback from users, and share feedback to the Informatics team? Can you explain and give examples?

**Section 3**: SUIT program screening

Q4: What has changed in the universal screening process since the adoption of the smart AI tool? How? Who reviews the universal screenings (U.S.) results, floor nurses or SUIT social workers? Where do you document about these results?

% of U.S. reviewed and/or administered by SW?

% of U.S. reviewed and/or administered by floor nurses?

Anyone else reviews or administers U.S., and if so, who and what % of the U.S.?

Q5: What has changed in process of administering the secondary screen (AUDIT/DAST) process since the adoption of the AI tool? How?

Q6: What has changed in the process of conducting the Brief Intervention/MI (mild/moderate condition vs. severe condition), Inpatient SUIT Consults, Inpatient MATs, and outpatient SUIT consults since the adoption of the AI tool? How?

***<u>GUIDE 3</u>***: *Brief Interview guide of SUIT program Social Workers and floor staff [*Unit (n=2) and ICU (n=3) registered nurses (RN)s, and unit SWs (n=4) as well as the SUIT SW (n=1)] – *not all questions may apply to all interviewees.*

Q1: What is your main role? Please pick one from the options below:

1. Emergency department nurse
2. Floor nurse
3. Floor social worker
4. SUIT social worker
5. Other?

<u>Section 1: Two substance use questions screening – Done by ED and floor nurse</u>

Q2: Do you usually conduct universal substance misuse screening by asking patients if they have misused alcohol and/or drug in the past year?

1. Yes
2. No – Please skip to Section 2, Question 5.

Q3: Can you explain in few words how you identify patients for universal screen? For example, which information you review to identify the patient and how long, on average, does it take you to identify a patient for universal screen?

Q4: On average, how long does it take you to administer a universal screen? Which factors, in your opinion, may impact the length of administering the universal screening?

Section 2: Secondary Screen by AUDIT/DAST questionnaires – Done by social workers (rarely ED and mostly floor social workers).

Q5: Do you usually conduct secondary screening by administering AUDIT/DAST questionnaires?

1. Yes
2. No – Thank you for your time. You have completed the brief interview.

Q6: Can you explain in few words how you identify patients for AUDIT/DAST screening? For example, which information you review to identify the patient and how long, on average, does it take you to identify a patient?

Q7: On average, how long does it take you to administer the AUDIT/DAST? Which factors, in your opinion, may impact the length of administering theAUDIT/DAST? ***Please do not account for the time it takes to deliver Brief Intervention/ Motivational Interviewing (BI/MI) which is the topic of the next section.***

Section 3: Brief Intervention/ Motivational Interviewing (BI/MI)

Q8: Do you usually conduct Brief Intervention/ Motivational Interviewing (BI/MI)?

1. Yes
2. No – Thank you for your time. You have completed the survey.

Q9: On average, how long does it take you to deliver Brief Intervention/Motivational Interviewing? What are the components, which factors, in your opinion, may impact the length of administering BI/MI?

### eFigure 1. Documentation Templates and Flow Sheets

Documentation templates and flow sheets were updated in the electronic health record (Epic Systems, Verona, WI) before the study began to standardize recording of screening methods, consultation reasons, intervention types, and patient outcomes. Templates included structured fields for: (1) screening method (manual nurse screening, manual social work screening, or SMART-AI notification); (2) substance type (alcohol, opioid, non-opioid drug, or polysubstance); (3) intervention delivered (brief intervention, motivational interviewing, naloxone dispensing, medication-assisted treatment initiation or adjustment, or outpatient referral); (4) patient response to intervention; and (5) follow-up plan.

### eFigure 1. Documentation Templates and Flow Sheets

| | | |
|---|---|---|
| **Substance Use Screen** | | |
| [[ WOMEN ]] How many times in the past year have you had 4 or… | None | |
| How many times in the past year have you used a recreational d… | 1 or more | |
| Excluding cannabis, how many times in the past year have you u… | | |
| **Alcohol Use Disorders Identification Test** | | |
| Frequency of Drinks Containing Alcohol | | |
| Number of Alcoholic Drinks on a Typical Day Drinking | | |
| Six or More Drinks on One Occasion | | |
| Unable to Stop Drinking | | |
| Fail to Perform Normal Expectations | | |
| Alcoholic Drink in the Morning After a Heavy Drinking Session | | |
| Guilt or Remorse After Drinking | | |
| Unable to Remember What Happened the Night Before Drinking | | |
| Injury as a Result of Your Drinking | | |
| Others Concerned About Your Drinking | | |
| Alcohol Use Disorders Identification Test | | |
| **Drug Abuse Screen Test** | | |
| Drugs used? | | narcotics (heroi… |
| How often have you used these drugs? | | Daily or almost … |
| Have you used drugs other than those required for medical reas… | | 0 |
| Do you abuse more than one drug at a time? | | 0 |
| Are you unable to stop using drugs when you want to? | | 1 |
| Have you ever had blackouts or flashbacks as a result of drug us… | | 0 |
| Do you ever feel bad or guilty about your drug use? | | 1 |
| Does your spouse (or parents) ever complain about your involve… | | 0 |
| Have you neglected your family because of your use of drugs? | | 0 |
| Have you engaged in illegal activities in order to obtain drugs? | | 0 |
| Have you ever experienced withdrawal symptoms (felt sick) whe… | | 1 |
| Have you had medical problems as a result of your drug use (e.g… | | 1 |
| Drug Abuse Screen Test | | 4 |

| | |
|---|---|
| **Floor SW Intervention** | |
| SW Intervention Completed? | |
| Recommended to SUIT Team | |
| Other Referrals | |
| **SUIT Intervention** | |
| Treatment | |
| Naloxone Counseling | |
| SUIT Consult Complete | |
| **SUIT Intervention and Outcomes** | |
| Consult Origin | Primary inpatie… |
| Consult Reason | Evaluation for a… |
| Screen Exemption | |
| Naloxone | |
| Medication-Assisted Treatment | |
| SUIT Social Work Intervention | |
| SUIT Provider Role | |
| Result of Consult Order | Defer to psychi… |
